# Endotracheal application of ultraviolet A light in critically ill severe acute respiratory syndrome coronavirus-2 patients: A first-in-human study

**DOI:** 10.1101/2021.03.05.21252997

**Authors:** Ali Rezaie, Gil Y. Melmed, Gabriela Leite, Ruchi Mathur, Will Takakura, Isabel Pedraza, Michael Lewis, Rekha Murthy, George Chaux, Mark Pimentel

**Affiliations:** Medically Associated Science and Technology (MAST) Program, Cedars-Sinai, Los Angeles; Karsh Division of Gastroenterology and Hepatology, Cedars-Sinai, Los Angeles; Division of Endocrinology, Diabetes and Metabolism, Department of Medicine, Cedars-Sinai, Los Angeles; Division of General Internal Medicine, Department of Medicine, Cedars-Sinai, Los Angeles; Division of Pulmonary and Critical Care Medicine, Department of Medicine, Cedars-Sinai, Los Angeles; Division of Infectious Diseases, Department of Medicine, Cedars-Sinai, Los Angeles

**Keywords:** SARS-CoV-2, ultraviolet-A light, endotracheal administration, viral load

## Abstract

**Background:** Promising preclinical experiments show that, under specific and monitored conditions, ultraviolet-A (UVA) exposure reduces certain bacteria, fungi, and viruses including coronavirus-229E without harming mammalian columnar epithelial cells. We aimed to evaluate the safety and effects of UVA therapy administered by a novel device via endotracheal tube in critically ill subjects with SARS-CoV-2 infection.

**Methods:** Five newly intubated mechanically ventilated adults with SARS-CoV-2 infection, with an endotracheal tube size 7.5mm or greater, were treated with UVA for 20 minutes daily for 5 days, and followed for 30 days.

**Results:** Five subjects were enrolled (mean age 56.6yrs, 3 male). At baseline, all subjects scored 9/10 on the WHO clinical severity scale (10=death) with predicted mortality ranging from 21 to 95%. Average log changes in endotracheal viral load from baseline to day 5 and day 6 were -2.41 (range -1.16 to -4.54; Friedman P=0.002) and -3.20 (range -1.20 to -6.77; Friedman P<0.001), respectively. There were no treatment-emergent adverse events. One subject died 17 days after enrollment due to intracranial hemorrhagic complications of anticoagulation while receiving extracorporeal membrane oxygenation. The remaining subjects clinically improved and scored 2, 4, 5, and 7 on the WHO scale at day 30. In these subjects, the slope of viral load reduction during UVA treatment correlated with the slope of improvement in clinical WHO severity score over time (Spearman rho=1, P<0.001).

**Conclusion:** In this first-in-human study, endotracheal UVA therapy under specific and monitored settings, was safe with a significant reduction in respiratory SARS-CoV-2 viral burden over the treatment period.

**Trial Registration:** ClinicalTrials.gov #NCT04572399.

**Key Messages:**

- What is the key question? Can endotracheal narrow-band UVA therapy be a safe and effective treatment for severe SARS-CoV-2 infection?
- What is the bottom line? Under specific and monitored settings, endotracheal UVA light therapy may be an effective treatment for SARS-CoV-2 infection. Endotracheal UVA light therapy appears to be well tolerated in critically ill patients with SARS-CoV-2 infection.
- Why read on? This is the fist-in-human trial of internal UVA therapy using a alternative novel approach to combat COVID-19.

## Introduction

Since the first report of severe acute respiratory syndrome coronavirus-2 (SARS-CoV-2) infection in December 2019, the global quest to find a highly effective modality to treat severe coronavirus disease 2019 (COVID-19) has been disappointing. Direct cytotoxic effects of SARS-CoV-2 along with dysregulated inflammatory responses and secondary respiratory infections continue to inflict substantial morbidity and mortality in severe and critical cases of COVID-19.[1-3] One mechanism to explain SARS-CoV-2 virulence may be through impairment of the mitochondrial antiviral signaling (MAVS) protein, responsible for innate antiviral responses.[4] Externally applied ultraviolet light therapy is an approved treatment for several atopic, inflammatory and dysplastic dermatologic disorders.[5] In preclinical experiments, ultraviolet-A (UVA) exposure under monitored conditions (i.e. specific intensity, peak wavelength, exposure time and distance to target tissue), reduces bacteria, fungi, and viruses including coronavirus-229E but does not harm human (*in vitro)* or murine (*in vivo)* columnar epithelial cells.[6] Moreover, UVA exposure of coronavirus-229E transfected human primary tracheal cells led to activation of the MAVS protein, reduction in spike protein, and resumption of cell proliferation similar to uninfected cells, suggesting that UVA may induce a beneficial antiviral state in infected human cells.[6]

Using these newly discovered principles, we aimed to investigate the safety and treatment effects of a novel device inserted into the endotracheal tube to deliver UVA and reduce the viral burden of respiratory SARS-CoV-2 in critically ill subjects.

## Methods

### Trial design

In this first-in-human study, we aimed to recruit and treat 5 subjects. The trial protocol (ClinicalTrials.gov number NCT04572399) was approved by the institutional review board of Cedars-Sinai, Los Angeles and was overseen by an independent data and safety monitoring board (DSMB). Subjects’ legally authorized representatives provided written informed consent. Inclusion criteria included age over 18 years, positive PCR test result for SARS-CoV-2 on nasal swab, and mechanical ventilation with an endotracheal tube (ETT) inner diameter of ≥7.5 mm. Pregnant women were excluded. Subjects received all standard supportive care; concomitant use of any other COVID-19 treatments was permitted.

### UVA device

The UVA therapy device (Aytu Biosciences, Englewood, CO) consisted of a 5.4 mm diameter sterile sealed multi-LED UVA light catheter within a protective sheath and endotracheal adaptor, umbilical, and control unit (Fig. S1). The UVA catheter adaptor was connected to the ETT using a double-swivel multi-access port (Halyard Health, GA) to maintain a closed-loop system and prevent ambient exposure to exhaled air upon introduction of the catheter into the ETT.

### Procedure

Within 24 hours of enrollment, subjects underwent 20 minutes of endotracheal UVA therapy, which was repeated once daily for a total of 5 consecutive days. All subjects received 100% FiO_2_ for 30 minutes prior to the procedure (see Supplemental Materials and Methods for protocol). The UVA catheter was inserted to the distal end of the ETT, with concomitant ventilator adjustments to flow rate and tidal volume to maintain optimal oxygenation. A plastic clamp fixed the catheter base to the access port to assure stability and consistent depth of catheter insertion throughout the 20-minute treatment session. The procedural instructional video can be accessed at: https://cedars.box.com/s/0lqm2slw1vlyt4j5em70xregfro4592t. UVA dosing was chosen based on the optimal response of coronavirus 229E-infected human primary tracheal cells to UVA exposure observed in *in vitro* experiments.[6] Controlled narrow-band UVA emission (peak wavelength 340-345nm) of maximum 2 milliwatt/cm^2^ was delivered at the level of tracheal mucosa. Predetermined criteria for treatment cessation and withdrawal of the UVA catheter included O_2_ saturation drop below 88% or hemodynamic instability.

Endotracheal (ET) aspirates were taken prior to each UVA treatment and 24 hours after the last UVA treatment for assessment of SARS-CoV-2 and absolute bacterial loads. Steps in preparation of the sampling traps and tracheal sampling, as well as sample processing and analysis for viral and absolute bacterial loads are provided in the Supplemental Materials and Methods. Absolute quantification of bacterial load represented culturable and non-culturable, viable and non-viable, pathogenic, and non-pathogenic bacteria.

Baseline, hospital, and ICU admission-related information including relevant clinical, laboratory, and radiologic data were recorded for all patients until 30 days after enrollment. The World Health Organization (WHO) COVID-19 10-point ordinal severity scale[7] was calculated at enrollment, and on days 15 and 30 following enrollment. SOFA[8] and SAPSIII[9,10] scores were calculated from the worst values within 24 hours of ICU admission.

### Outcomes and statistical analysis

The primary endpoint was the change in ET aspirate SARS-CoV-2 viral load from day 0 to the last day of treatment. Secondary outcomes included treatment-emergent adverse events (TEAEs), changes in endotracheal absolute bacterial load, clinical outcomes and laboratory parameters including inflammatory markers, and changes in the WHO COVID-19 10-point ordinal scale of improvement from baseline to day 15 and 30.

Freidman test was used to detect differences across daily viral and bacterial loads. One sample t-test was used to analyze changes in inflammatory markers and microbial loads from day 0 to day 1[11]. Spearman rank-order test was used to assess correlations between the reduction of viral load (log_10_) and the improvement of WHO scale. The reduction of viral load (log) from baseline to the final endotracheal sample was defined as the slope of the linear regression between log_10_ viral load and time point of viral load measurements. Similarly, the estimated improvement of WHO scale from baseline through day 30 was the slope of the linear regression between WHO scale and the time of WHO scale measurements. A significance level of α=0.05 was used.

## Results

Between October 30, 2020 and November 28, 2020, 5 subjects were enrolled (mean age 56.6yrs, 3 male). Baseline characteristics of the enrolled subjects are summarized in Table 1, and a summary of the timeline and key events is presented in Figure 1. At the time of intubation, all 5 patients were critically ill, with WHO COVID-19 ordinal scale scores of 9 in all subjects, and with SOFA scores predicting a 20-95% mortality rate. All patients received daily 20-minute treatments starting within the first 36 hours following intubation, for 5 days. Baseline and day 6 ET aspirates were taken in all patients except for study subject #1 who was extubated on day 6. Hence, a total of 29 ET aspirates were analyzed.

**Table 1.** Baseline characteristics on the day of intubation.

| **Subject** | **1** | **2** | **3** | **4** | **5** |
| --- | --- | --- | --- | --- | --- |
| **Age** | 65 | 38 | 64 | 62 | 54 |
| **Gender** | M | M | M | F | F |
| **Race/ Ethnicity** | White/<br>Hispanic | White/<br>Hispanic | White/<br>Persian | African<br>American | White/<br>Hispanic |
| **BMI** | 26.0 | 36.3 | 25.5 | 35.4 | 34.0 |
| **PMH** | Type 2 DM | Prediabetes | Type 2<br>DM, HTN | Mechanical<br>mitral valve,<br>HTN,<br>Dyslipidemia | Type 2 DM |
| **Symptom onset to intubation (days)** | 14 | 18 | 11 | 5 | 10 |
| **ETT size (mm)** | 7.5 | 8.0 | 8.0 | 7.5 | 7.5 |
| **PaO<sub>2</sub>/FiO<sub>2</sub>** | 70 | 51 | 50 | 50 | 82 |
| **Vasopressor use** | + | + | + | + | + |
| **ECMO** | - | + | - | - | - |
| **SOFA score/predicted mortality** | 8/33.3% | 8/33.3% * | 14/95.2% | 8/33.3% | 7/21.5% |
| **SAPSIII score/predicted mortality** | 62/34% | 62/34% * | 85/67% | 68/43% | 57/26% |

\*Note that SOFA and SAPSIII scores do not account for the need for ECMO
BMI, Body mass index; ECMO, Extracorporeal membrane oxygenation, DM, Diabetes mellitus; ETT, Endotracheal tube; HTN, Hypertension; PMH, Past medical history; SAPSIII, Simplified Acute Physiology Score III; SOFA, Sequential organ failure assessment.

**Figure 1.**
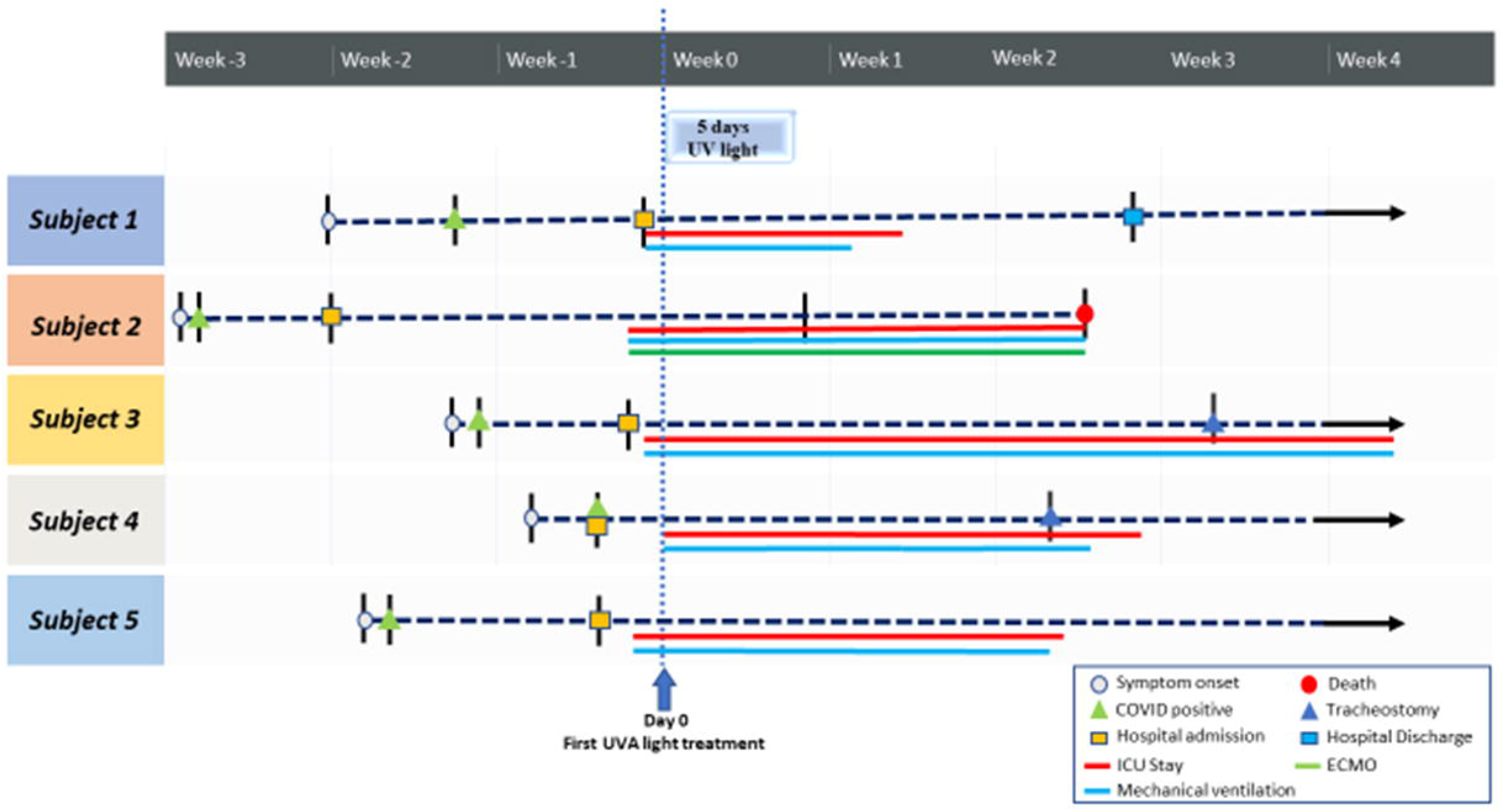
Summary of the timeline and key events for enrolled subjects.

### Primary Outcome

Subjects had elevated viral loads at baseline (range 3.4×10^4^ -1.64×10^7^ copies/ml) except for study subject 2 who had an undetectable viral load at all time points, demonstrating that virus had cleared since the last nasal swab (Fig. S2). There was no significant correlation between symptom onset date and either baseline (Spearman rho=-0.70, p=0.23) or day 6 viral loads (Spearman rho=-0.21, p=0.83).

There was a significant reduction of SARS-CoV-2 levels in endotracheal aspirates during UVA treatment. The average log_10_ changes in endotracheal viral load from baseline to day 5 and day 6 were -2.41 (range -1.16 to -4.54; Friedman p=0.002) and -3.2 (range -1.2 to -6.77; Friedman p<0.001), respectively (Fig. 2, Fig. S2).

**Figure 2.**
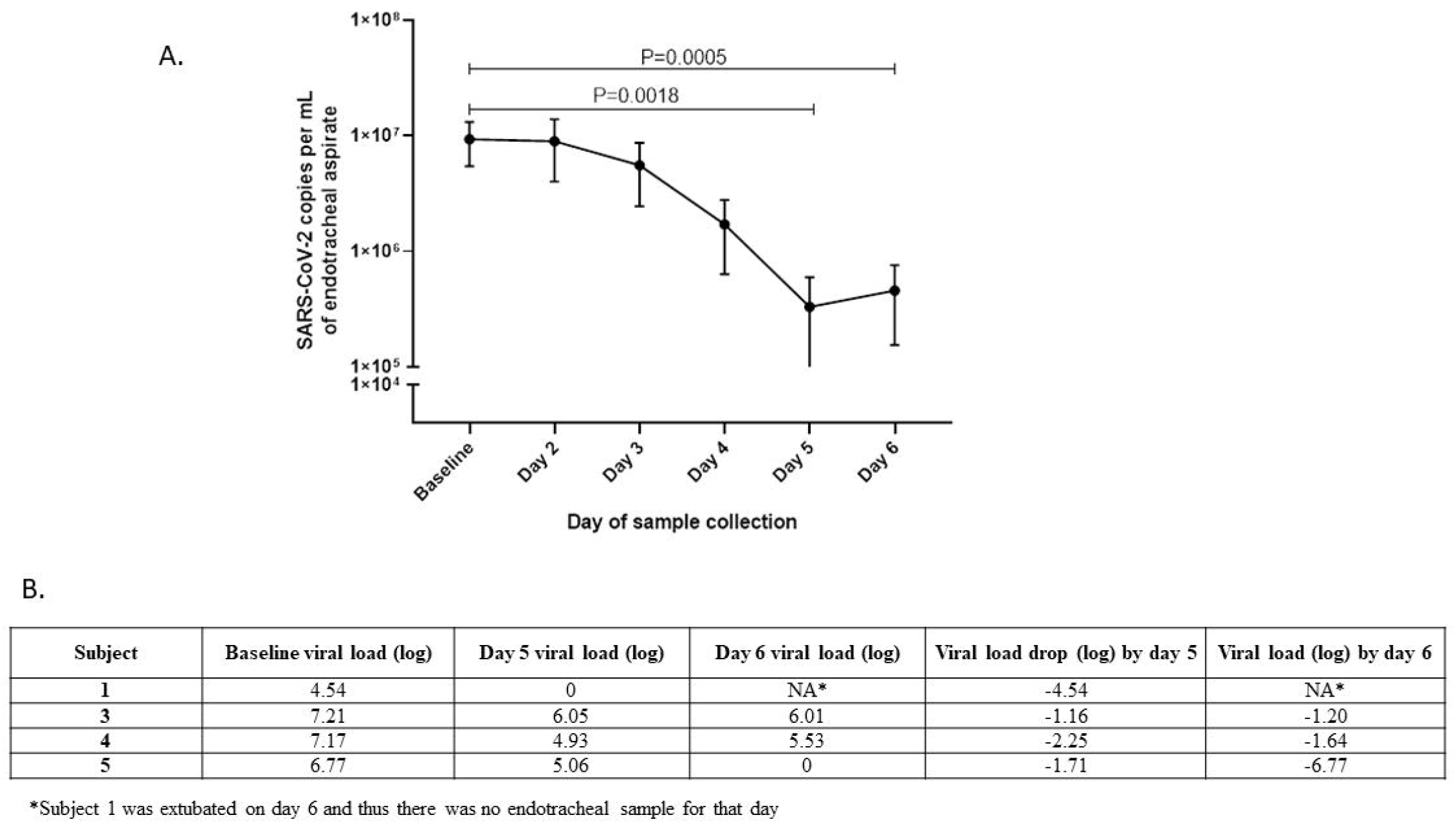
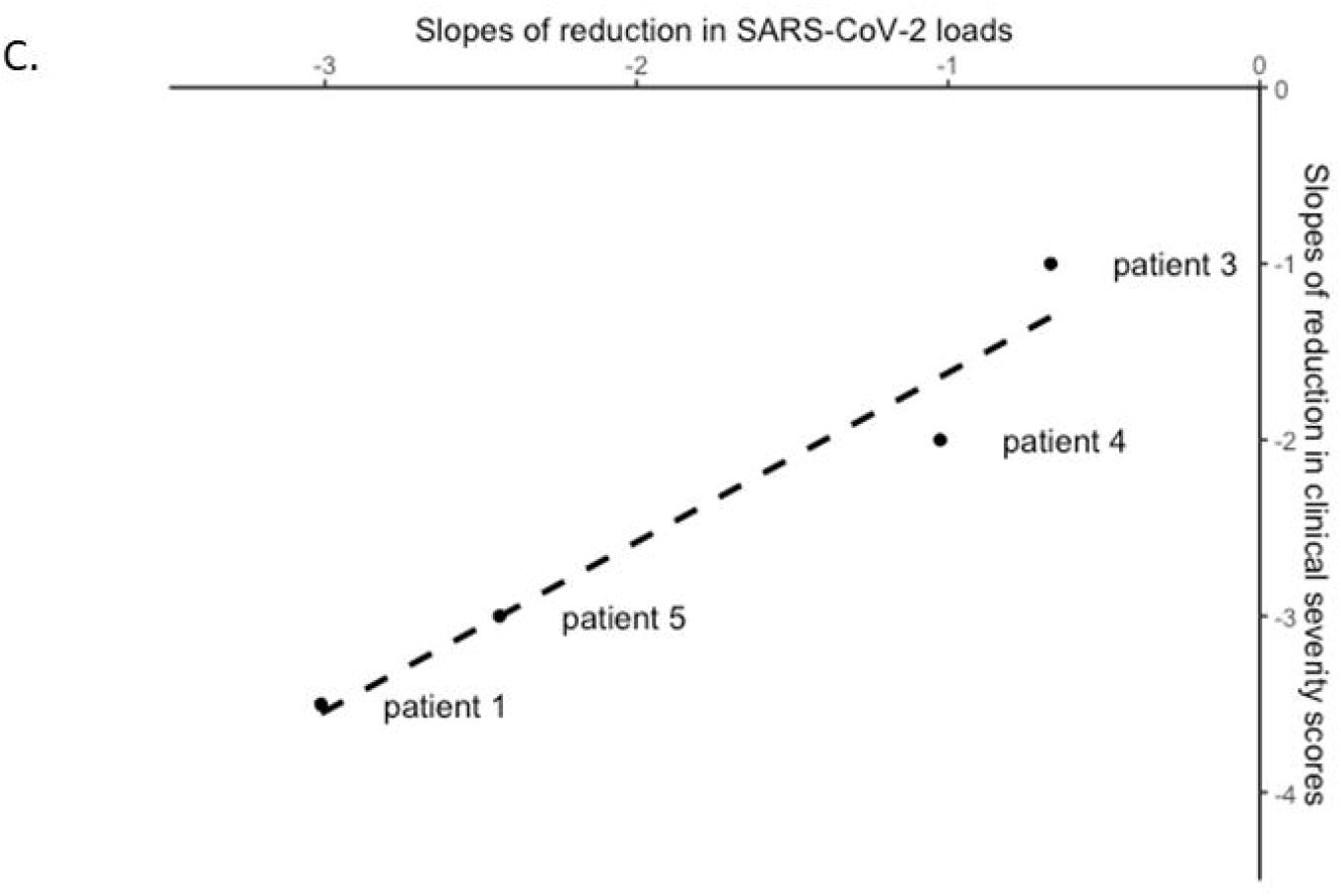
A) Reductions in endotracheal SARS-COV-2 loads from day 0 through day 6 in patients with detectable viral load at baseline. Freidman test is used to analyze differences across daily viral load measurements. B) Corresponding viral loads (log) for each subject at baseline, day 5 and 6. Average log changes from baseline to day 5 and day 6 were -2.41(relative reduction >99%) and -3.2 (relative reduction >99.9%), respectively. C) The individual slopes of reduction in SARS-CoV-2 loads during UVA therapy correlated with the slopes of reduction in WHO severity score by day 30.

### Secondary and Clinical Outcomes

Among the secondary outcome measures, quantification of absolute endotracheal bacterial load at baseline ranged from 1×10^3^ -1.7x 10^6^ CFU/ml and remained statistically unchanged during the UVA treatment sessions (Fig. S3).

The clinical course for each subject is shown in Figure 1. WHO clinical severity scores improved by an average of 1.6 and 3.6 points on day 15 and day 30, respectively. Excluding subject 2 who had undetectable baseline viral load, WHO severity scores improved by 4.75 points on day 30 (Table S1). All subjects survived except study subject 2, who was placed on comfort care following intracranial hemorrhage due to ECMO-associated anticoagulation and died on ICU day 17. Interestingly, there was an association between WHO clinical severity score outcomes and viral reductions during UVA treatment. Improvement in WHO severity scores by day 30 exhibited a positive correlation with the reduction of viral load during UVA therapy (Spearman’s rho=1, p<0.001) (Fig. 2c). Following UVA therapy, there was a significant drop in C-reactive protein (-95.0 ± 48.0 mg/L, p=0.04) within 7 days of enrollment. Observed reductions in interleukin-6 (-258.9 ± 621.4 pg/mL, p=047) and ferritin (-563.6 ± 514.8 ng/mL, p=0.12) did not reach statistical significance (Table S2).

### Safety outcomes

No treatment-emergent adverse events or need for treatment cessation was observed in the study. Oxygen saturations and hemodynamics during all treatment sessions remained stable. None of the subjects experienced pneumothorax, subcutaneous emphysema, or endotracheal tube (ETT) dislodgment. Adverse events were deemed unrelated to UVA therapy (Table S3). Two subjects eventually underwent bronchoscopy for tracheostomy tube placement for prolonged intubation which revealed normal looking tracheae without erythema or friability (Fig. S4). An independent Data and Safety Monitoring Board did not recommend any changes to the treatment protocol for future planned trials.

## Discussion

The global challenge associated with the COVID-19 pandemic is a bitter reminder that safe and effective therapies are desperately needed to treat resistant and/or novel pathogens. While externally applied UV therapy is commonly used in dermatologic diseases, due to technological limitations and knowledge gaps, internal UV therapy has never previously been performed. In this first-in-human study, endotracheal UVA light appeared safe in critically ill patients with COVID-19. Furthermore, a significant reduction in endotracheal SARS-CoV-2 levels was observed following 5 days of UVA therapy. Finally, the reduction of viral load during UVA treatment correlated with the reduction in the WHO clinical severity scores.

Apart from a dysregulated inflammatory response, heightened viral replication has a critical role in pathogenesis of severe and critical COVID-19. There is a significant association between respiratory SARS-CoV-2 load and mortality.[12] In addition, severe cases of COVID-19 exhibit longer duration and a later peak of virus in respiratory samples as compared to mild disease.[13] Aligned with these findings, 4 out of 5 of our subjects had high viral loads in the endotracheal aspirate at baseline ICU care without a significant correlation with the time of symptom onset. In addition, improvement of WHO clinical severity scores by day 30 significantly correlated with the reduction of viral load during UVA therapy. Taken together, these suggest a temporal overlap between the viral replication and hyperinflammatory phases[2] in the disease course of critically ill COVID-19 patients. Hence, these patients may continue to benefit both from viral load reduction and from improvement of the innate immune response to SARS-CoV-2. UVA potentially possesses the ability to provide benefit in both areas. Firstly, we have shown previously that UVA therapy exhibits antiviral effects against positive sense, single-stranded RNA viruses including coxsackievirus and coronavirus-229E.[6] Secondly, *in vitro* UVA exposure led to activation of MAVS protein in virally infected primary human tracheal cells, a pathway that is directly impaired by SARS-CoV-2. Activation of the MAVS protein pathway by UVA exposure in SARS-CoV-2-infected tracheal cells may be a mechanism behind the significant reduction of viral load along the respiratory tract in our study despite only intermittent and localized UVA therapy in the upper airway.

As compared to conventional microbial cultures, absolute bacterial load quantified by PCR detects a greater number of bacteria including unculturable and non-viable bacteria yielding higher load of bacteria. We did not detect a significant change in endotracheal aspirate bacterial loads during UV therapy. This is encouraging and is likely due to leveraging a closed-loop system for introduction of the sterile UVA catheter. We previously have shown that UVA reduces several known pathogens linked to ventilator associated pneumonia (VAP) including *Pseudomonas aeruginosa, Klebsiella pneumoniae, Escherichia coli, Enterococcus faecalis, Streptococcus pyogenes, Staphylococcus epidermidis* and *Candida albicans*.[14] The potential role of UVA therapy in the prevention of VAP by decreasing or delaying tracheal and ETT colonization of pathogenic bacteria warrants further assessment but the lack of rise in bacteria seen here is promising. No treatment emergent adverse events occurred during the 25 UVA treatment sessions and serious/severe adverse events were unrelated to the treatment intervention. Oxygenation and hemodynamics remained stable during all treatments. Subsequent bronchoscopy in 2 subjects revealed normal-looking trachea, consistent with our preclinical *in-vivo* and *in-vitro* safety experiments.[6] Subject number 2 died due to complications of ECMO-related anticoagulation (intracranial hemorrhage) despite stable oxygenation at the time of stroke. Bleeding occurs in approximately 50% of patients undergoing ECMO [15] with intracranial hemorrhage having an 85% risk of mortality.[16] Despite being in a highly critical state, 4 out of 5 subjects survived and had meaningful clinical improvements (Table S1). Further trials are needed to elucidate whether UVA therapy can improve clinical outcomes.

Our study has several limitations. As this was a first-in-human trial, the sample size was small. However, subjects had a diverse distribution of several known risk factors for severity of COVID-19 including age (range 38-65 years), sex (2 females and 3 males), race (1 non-Hispanic white, 3 Hispanic White and 1 African American) and BMI (range 25-36). Of 5 patients, 3 had the smallest allowable ETT size (7.5 mm) without any treatment-emergent adverse events. With rapid advancements in LED and fiberoptic technology, future designs may accommodate patients with smaller diameter ETTs. Finally, the natural history of SARS-CoV-2 load in endotracheal aspirates is poorly defined. Zheng et al. observed a mean baseline respiratory viral load of 10^5^ copies/ml in 74 severe cases and a very gradual rate of viral clearance in lower respiratory tract (27.7 days from onset of symptoms) in 29 patients admitted to ICU.[13] The 3.2 log reduction in our study after 5 days of UVA therapy appears to outpace the natural decline of respiratory viral load. Further study may help characterize the natural history of SARS-CoV-2 levels in the respiratory tract of ICU subjects.

Using a novel device in a specific and monitored setting, endotracheal UVA therapy in critically ill subjects is associated with a significant reduction of respiratory SARS-CoV-2 viral load. Viral load reduction correlated with improvements in the WHO severity score by day 30. Finally, to date there does not appear to be any treatment-emergent adverse outcomes from the direct effects of the UVA or the mechanical effects of endotracheal catheter insertion.

## Data Availability

All data are in the manuscript and supporting files

## Acknowledgments

The authors sincerely thank: Shane White, Melissa Hampton, Elizabeth Khanishian, Christine Chang, Peter Chen, and Dr. Gillian Barlow for their assistance in the conduct of the study; Jiajing Wang for statistical analysis; the Cedars Sinai respiratory therapy team including LaShone Mays and Brian Richards; the Cedars Sinai Women’s Guild Simulation Center and Michael Lappen for development of instructional video; and Sterling Medical Devices. We also thank Frank Lee for his support of the MAST Program’s COVID-19 research.

## Author contributions

AR, GYM, RM, MP, GC, IP, ML, RM designed the study; AR, GYM, WT, GL, GC, IP acquired the data; AR, GYM, GL, RM, MP, WT, GC analyzed and interpreted the data; all authors revised the manuscript for intellectual content.

## Availability of data and materials

All data is available in the main text or the supplementary materials.

