## Supplemental Materials for "Endotracheal application of ultraviolet A light in critically ill severe acute respiratory syndrome coronavirus-2 patients: A first-in-human study"

**Supplementary Materials**

**Supplemental Figures and Tables**

**Fig. S1.** Device components. Left panel shows the controller, umbilical (flexible power and air connector between the catheter and the controller) and the air compressor. Right panel shows the catheter enclosed in a plastic sleeve with a port that connects to double-swivel multi-access port. Chilled air flows inside the catheter during treatment and a heat-detecting thermistor will automatically shuts off the controller if catheter temperature rises. The catheter can be safely withdrawn from the endotracheal tube back into the plastic sleeve until reuse in 24 hrs.

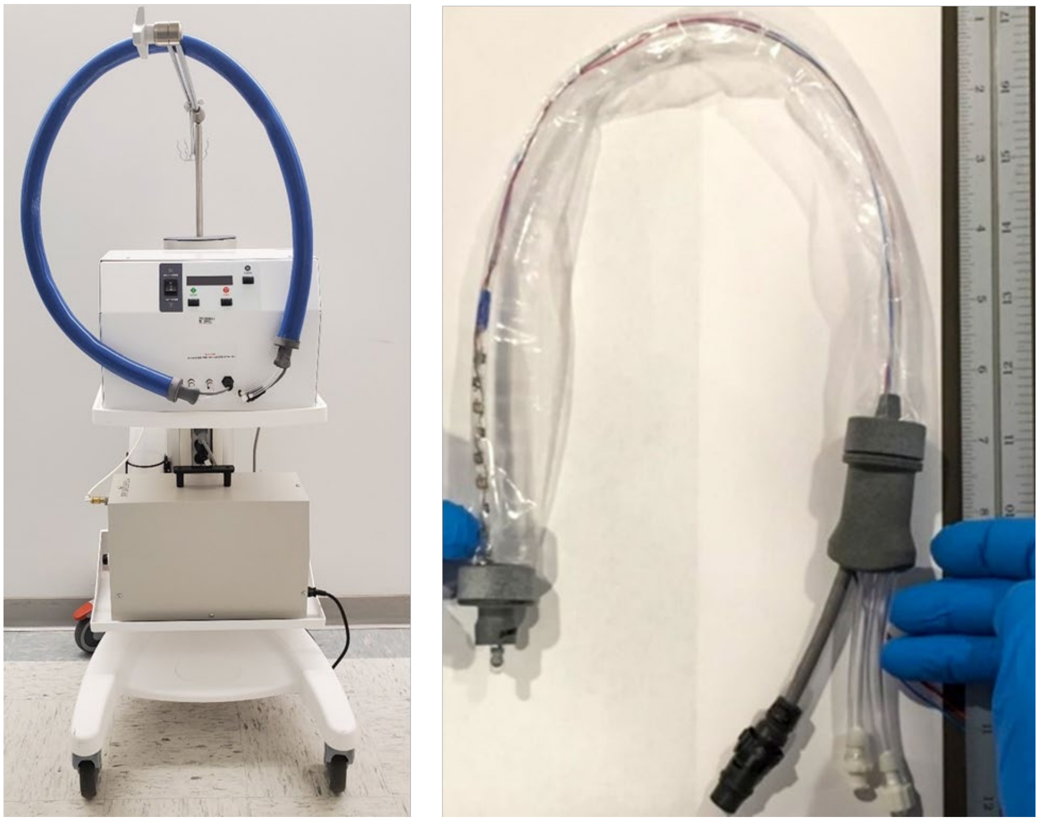

**Fig. S2.** Individual daily quantification of endotracheal SARS-CoV-2 loads. Subject 2 did not have detectable viral load at any point of the study.

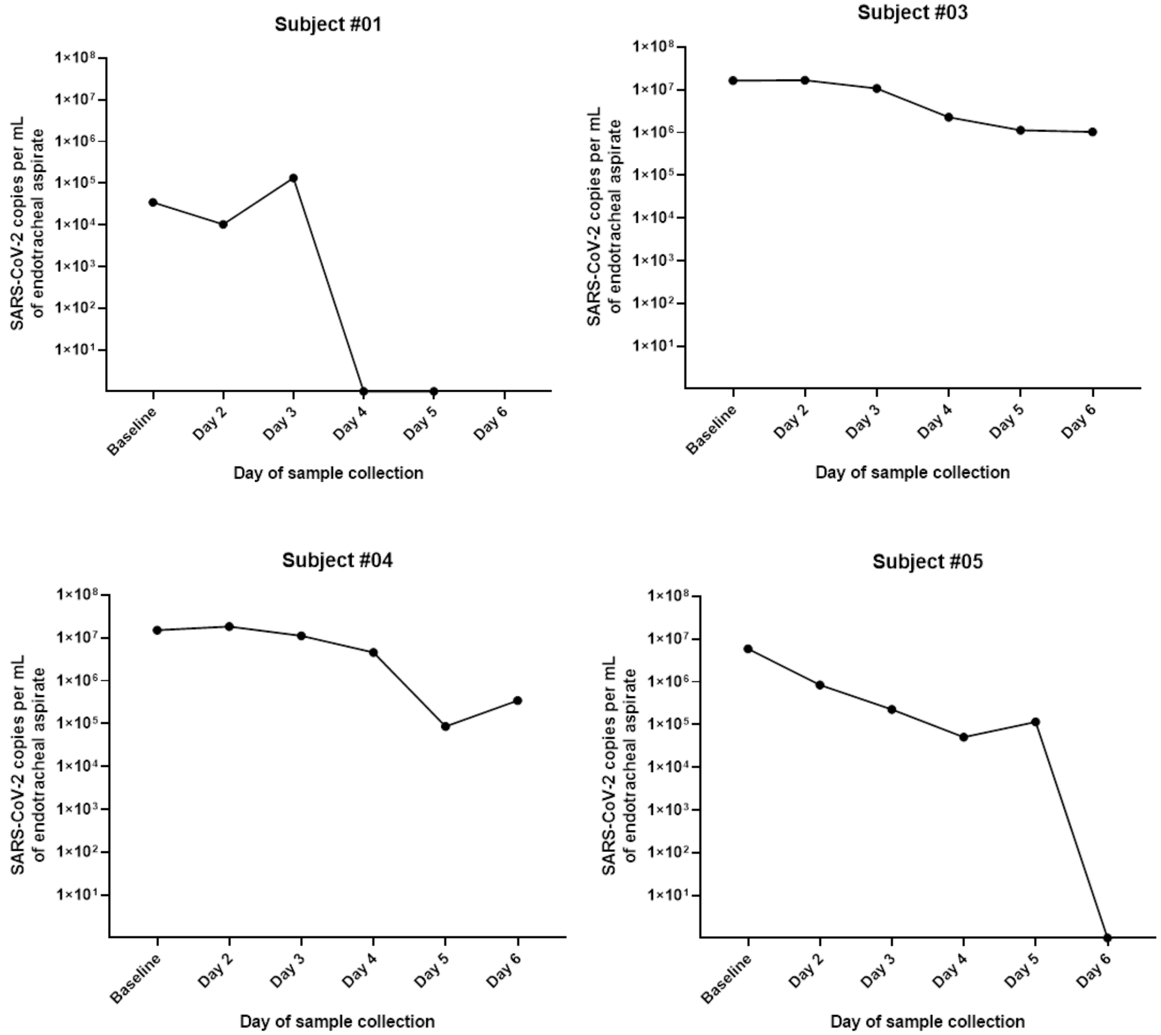

**Fig. S3.** a) Average daily quantification of endotracheal absolute bacterial loads from day 0 to day 6. b) Individual daily quantification of endotracheal absolute bacterial loads from day 0 to day 6.

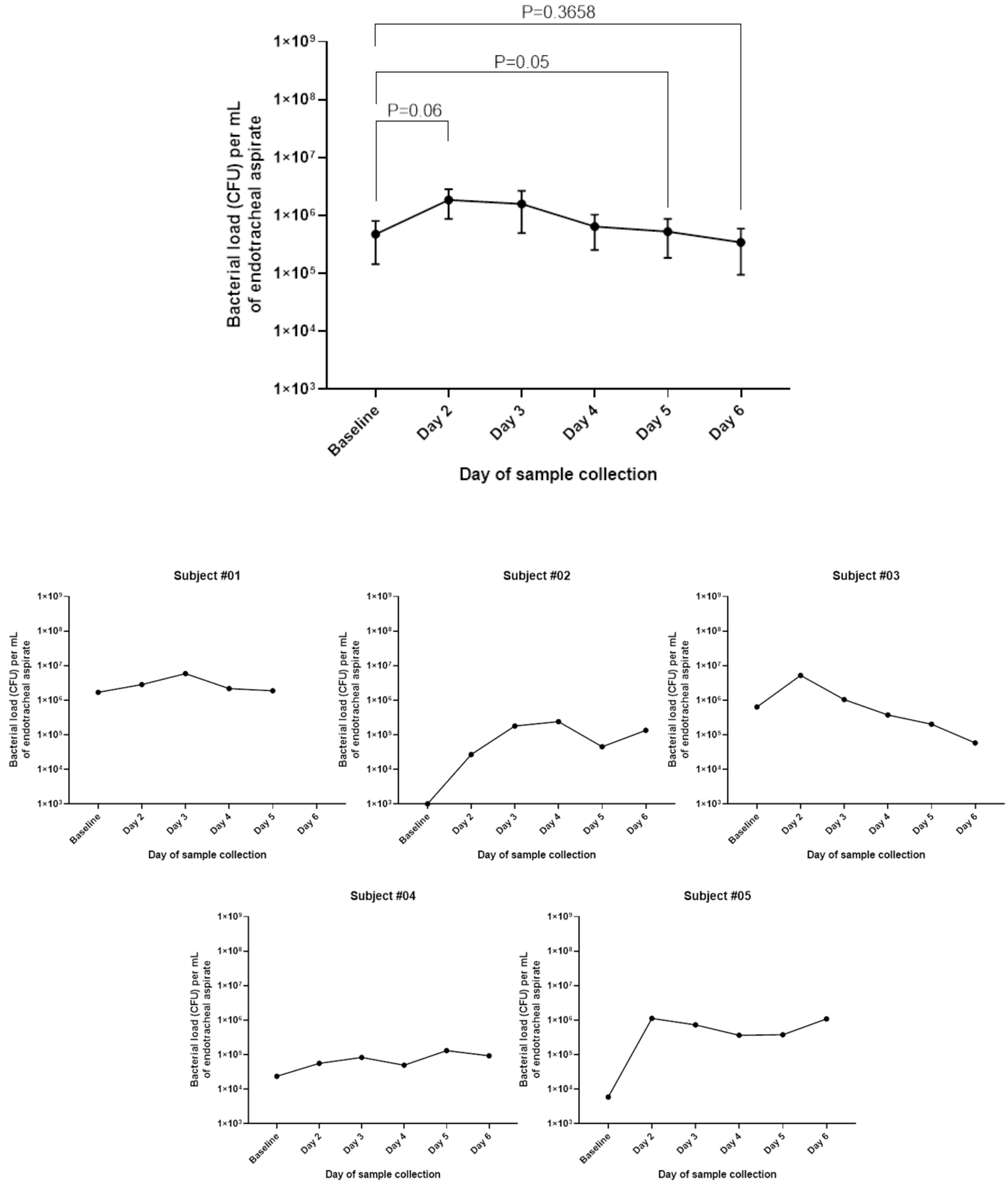

**Fig. S4.** Bronchoscopy pictures during tracheostomy for patient #3 showing normal trachea without erythema, edema or friability (left panel). Full thickness penetration of the tracheal wall did not lead to excessive bleeding or hematoma (right panel).

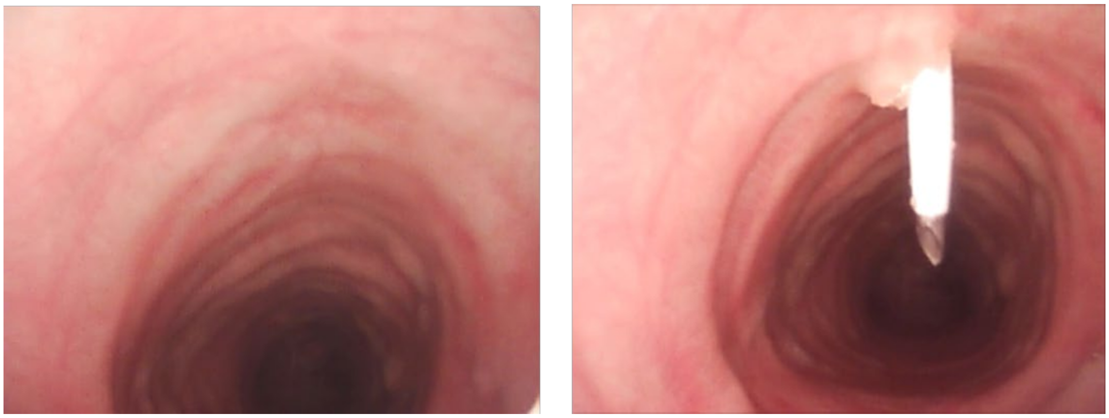

**Table S1.** World Health Organization clinical severity scale at baseline, day 15 and 30.

| Subject | Baseline | Day 15 | Day 30 | Subject status at day 30 |
|---|---|---|---|---|
| **1** | 9 | 5 | 2 | Independent at home. Occasional 1 lit/min $O_2$ when ambulating |
| **2*** | 9 | 9 | 10 | Fatal ECMO-related intracranial hemorrhage on day 17 |
| **3** | 9 | 9 | 7 | Ventilated, $FiO_2$ 50%, PEEP 5, $O_2$ Saturation 94% |
| **4** | 9 | 7 | 5 | Ambulating, 3 lit/min $O_2$ via tracheostomy collar |
| **5** | 9 | 7 | 3 | On room air. Needing assistance for ambulation |

**Table S2.** Change in inflammatory markers following UVA therapy.

| | Baseline (Mean ± SD) | Post UV therapy (Mean ± SD) | Change (Mean ± SD) | P value |
|---|---|---|---|---|
| **CRP (mg/L)** | 164.13 ± 87.24 | 80.2 ± 64.19 | -95.03 ± 47.97 | 0.04 |
| **Ferritin (ng/mL)** | 1273.18 ± 871.58 | 709.55 ± 439.77 | -563.63 ± 514.78 | 0.12 |
| **Interleukin-6 (pg/mL)** | 308.15 ± 585.34 | 49.25 ± 87.83 | -258.9 ± 621.40 | 0.47 |

Data was not available for one patient. CRP, C-reactive protein; SD, standard deviation

**Table S3.** Serious/severe adverse events from day 0 to day 30. None of the events were deemed related to UV therapy by the investigators or drug and safety monitoring board (DSMB). None of the adverse event occurred during or immediately after UVA treatment sessions.

| Subject | Serious/severe adverse events |
|---|---|
| **1** | *Veilonella* bacteremia and *Candida albicans* fungemia |
| **2** | *E. Fecalis* bacteremia, spontaneous hemorrhage from lines and cerebellar and intraventricular hemorrhage with ventriculomegaly due to ECMO-related anticoagulation |
| **3** | Acute renal failure post intravenous contrast requiring dialysis, stress cardiomyopathy requiring pulse steroids, thrombocytopenia, rapid ventricular response atrial fibrillation, congestive hepatitis, compartment syndrome of right forearm due to proning requiring fasciotomy, bleeding from forearm wound vac, *Staphylococcus capitis* bacteremia, *Clostridium difficile* colitis, rectal bleeding due to rectal tube pressure requiring transfusion |
| **4** | *Enterococcus Faecalis* bacteremia, vancomycin-associated face/tongue swelling; transient 30 second run of ventricular tachycardia, hematoma of the right arm, aspiration pneumonia post extubation |
| **5** | Probable ventilator associated pneumonia with purulent endotracheal secretions and positive endotracheal culture for *Pseudomonas aeruginosa* |

**Supplementary Materials and Methods**

A) Protocol for insertion of UVA catheter.

1. Ensure bite block is inserted
2. Ensure that patient has ETT size 7.5mm or greater, attached to the mutliaccess port
3. The device can be inserted with the patient in either the supine or the prone position
4. Ensure $FIO_2$ is at 100% for at least 30 minutes prior to device deployment
5. Patient should be ventilated in volume control mode similar to bronchscopy
6. Perform routine respiratory suctioning of the endotracheal tube to ensure no mucous plug inside the tube.
7. The procedure needs to be performed in a manner that minimizes the likelihood of breaking the ventilatory circuit and exposing the circuit contents to ambient surroundings. The catheter will be enclosed in a plastic sleeve with a port that connects to the Y-port in the ventilatory circuit at or near the patient's mouth.
8. One of the study investigators must be present during the time period that the catheter is inserted in ETT.
9. Inspect tubing and catheter device for damage.
10. By holding the tubing, advance the catheter device approximately into the ventilatory circuit and the endotracheal tube, towards the carina, until the catheter marking aligns with the endotracheal tube entrance. Apply the appropriate clamp based on the size of the ETT.
11. Check the Controller for the green light to turn on.
12. Turn on the compressor and the UV light using the Controller. The 20-minute timer will begin automatically.

Upon completion of the 20-minute treatment, the timer will be at zero, and the light will automatically shut off. The catheter can be safely withdrawn from the endotracheal tube back into the plastic sleeve until reuse in 24 hrs.

B) Protocols for preparation of the sampling traps prior to transfer to ICU and tracheal sampling.

1. Under the clean and UVC-decontaminated hood, unpack and open the trap a. The hood should be set up for the trap preparation with 1000 µL, 200 µL, 20 µL and 10 µL sterile tips with barrier, biohazard sharp container, biohazard autoclavable bags, rack for cryovials, Sani Wipe or Virex, tape, scissors and pen. The hood should not be overloaded. After preparation, the hood should be clean and sterilized by turning on the UVC lamp for 20 minutes.
2. Transfer 2cc of buffer ATL (Qiagen) to the trap
3. Transfer 0.5cc of DTT to the trap
4. Close the trap using the lid with connectors
5. Attach the subject information label
6. Place the closed trap along with the solid lid (to be used after sample collection) back in the package
7. Transfer to ICU

Prior to entering the patient's room ensure that the following items are present:
a. Specimen trap
b. Three biohazard bags (two bags will be taken into the room and one stays outside the room)

c. Marker
d. Proper Personal Protection Equipment as per institutional policy
e. Alcohol swabs
f. Transportation box

- Preparation
  - Don appropriate PPE as per institution's policy for patients with COVID-19
  - Attach the suction tubing to the wall socket
  - Attach control valve to wall suction tubing.
  - Set vacuum regulator to 80-120 mmHg negative pressure
  - Detach the control valve from wall suction tubing and apply alcohol swabs to the control valve and the tip of the suction tubing
  - Connect the specimen trap between the suction control valve and the suctioning tubing
  - Turn ventilator to 100% $FiO_2$

- Sampling/suction procedure
  - Advance catheter 2cm proximal to the distal tip of the ETT.
  - Depress the control valve to apply suction and gently withdraw the suction catheter
  - If desired amount of sputum is collected in the trap. Stop suctioning, withdraw the catheter into the sheath and remove the trap from the suction tubing and then control valve.
  - If desired amount of sputum is not collected, instill saline into the ETT in 5 cc increments and repeat the steps above.
  - Tightly seal the trap by placing the rubber tubing over the male adaptor and place into the biohazard bag.
  - Reconnect suction tubing directly to the control valve and continue suctioning procedure as per institutional policy.
  - Record date/time, amount of saline instilled in ETT, treatment session number, medical record number and study ID on the biohazard bag using the marker and place the bag into the second biohazard bag.
  - Doff the gown and first set of gloves inside the room.
  - Outside the room, disinfect the biohazard bag and follow the doffing steps as per institutional policy.
  - Don a new set of gloves.
  - Place the sample into the autoclavable transportation box to be transferred to the lab.

C) Endotracheal sample microbiologic processing steps.

Aspirates from ETT were collected in a pre-weighted 40 mL Argyle sputum trap device (Covidien, Dublin, Ireland) 4 mL of ATL lysis buffer (cat. n° 19076, Qiagen, Hilden, Germany).

All steps involving samples processing after collection were performed in a clean and sterile BSL-2 hood. After collection, sputum trap with sample was weighted and extra ATL buffer was added when needed (when more than 4 mL of aspirate were collected, adjusting for at least 1:1 of sample/ATL). At least 1 mL of 1 x SPUTOLYSIN® Reagent – DTT (CAS 578517, MilliporeSigma, Burlington, MA) was added to the samples. Extra DTT was added to samples which were thick and hard to liquefy. After adding DTT, samples were mixed by vortexing until the sputum was fully liquefied. Samples were left sitting for 5 to 10 minutes to reduce the amount of foam. The resulted solution (ETT aspirate + ATL + DTT) was aliquoted in sterile 2 mL cryovial and stored in -80°C.

SPUTOLYSIN® Reagent liquefies the sputum samples and ensures that the bacteria and virus trapped in the mucus are exposed to the lysis solution. The ATL buffer inactivates samples by disrupting all viral and microbial population (including COVID-19) and at the same time expose their DNA and RNA to be extracted later.

*RNA/DNA extraction*

Samples were extracted with the kit MagMAX™ Viral/Pathogen Nucleic Acid Isolation Kit (cat. n° A42352, Thermo Fisher Scientific, Waltham, MA) according to the manufacturer modified protocol[1] for total nucleic acid purification - large volume: 500 µL to 2 mL, in a 24 Deep-well plate (Thermo Fisher Scientific, Waltham, MA). The extractions were performed in a closed automated system: Thermo Scientific KingFisher Duo Prime (cat. n° 5400110, Thermo Fisher Scientific, Waltham, MA) previously sterilized with UVC for 20 minutes, programmed with the file MVP_DUO_LV.bdz[2].

At the time of DNA/RNA extraction, at least 2.5 mL of lysed samples (ETT aspirate + ATL + DTT) were thawed in ice and transferred to a KingFisher™ deep-well 24 plate in row A. 100 µL of Proteinase K was added to the sample well and mixed by pipetting up and down. 100 µL of total nucleic acid magnetic beads was added to each sample (row A) and mixed again. 2 mL of wash buffer (row B) and 80% ethanol (rows C and D) were added into the 24 deep-well plate sample plate and placed in the KingFisher Duo along with the elution plate - another 24 deep-well plate containing 150 µL of the elution buffer in the row A and tip comb in row B. KingFisher Duo Prime was previously programmed with the file MVP_DUO_LV.bdz. After the end of the extraction, the elution plate was removed from the machine and samples containing the extracted DNA/RNA were aliquoted and stored at -80°C.

*SARS-CoV-2 load analysis*

SARS-CoV-2 load were analyzed in RNA samples extracted from tracheal aspirates with the TaqMan 2019-nCoV Assay Kit v1 (cat. n° A47532, Thermo Fisher Scientific) and TaqPath 1-Step RT-qPCR Master Mix, CG (cat. n° A15299, Thermo Fisher Scientific) following manufacturer protocol[3] as it is, without modifications. A standard curve was prepared using the TaqMan 2019-nCoV Control Kit v1 (cat. n° A47533, Thermo Fisher Scientific). The reaction

---

[1] User guide: MagMAX Viral/Pathogen Nucleic Acid Isolation Kit High throughput isolation of viral nucleic acid (RNA and DNA) from biofluids and transport media (https://www.thermofisher.com/document-connect/document-connect.html?url=https%3A%2F%2Fassets.thermofisher.com%2FTFS-Assets%2FLSG%2Fmanuals%2FMAN0018073_MagMAXViralPathoNuclAcidIsolatKit_Automated_UG.pdf&title=VXNlciBHdWlkZTogTWFnTUFYIFZpcmFsL1BhdGhvZ2VuIE51Y2xlaWMgQWNpZCBJc29sYXRpb24gS2l0LSBIaWdoIHRocm91Z2hwdXQgaXNvbGF0aW9uIG9mIHZpcmFsIG51Y2xlaWMgYWNpZCAoUk5BIGFuZCBETkEpIGZyb20gYmlvZmx1aWRzIGFuZCB0cmFuc3BvcnQgbWVkaWE=). Page 10 “Perform total nucleic acid purification using KingFisher™ Duo Prime (large volume: 500 µL to 2 mL)”.

[2] Available at: https://www.thermofisher.com/order/catalog/product/A42352?SID=srch-hj-A42352#/A42352?SID=srch-hj-A42352 File name: KingFisher Duo Protocol: MagMAX Viral/Pathogen Nucleic Acid Isolation Kit under “Product Literature”.

[3] Leite G et al. Optimizing microbiome sequencing for small intestinal aspirates: validation of novel techniques through the REIMAGINE study. BMC Microbiol. 2019; 19: 239.

[3] Available at https://www.thermofisher.com/order/catalog/product/A47532#/A47532 under “Manuals and Protocols”, file: Product Sheet: TaqMan™ 2019-nCoV Assay Kit v1.pdf

was performed in a MicroAmp™ Optical 384-Well Reaction Plates (Thermo Fisher Scientific) in a QuantStudio™ 6 Flex Real-Time PCR System (Thermo Fisher Scientific). Three SARS-CoV-2 genes were analyzed: ORF1ab, S protein and N protein.
The resulted Ct values were first classified and interpreted (Tables 1 and 2), then SARS-CoV-2 loads were calculated by comparing the Ct of positive samples with the Ct of the standard curve created with the TaqMan 2019-nCoV Control Kit v1, which had 50 µL (1 x $10^4$ copies/µL) and adjusted by dilutions and volume used for sample extraction.

Table 1. Classification of the SARS-CoV-2 results for each gene analyzed, according to the Ct values

| 2019-nCoV assay (FAM™ dye) | RNAseP assay (IPC) (VIC™ dye) | 2019-nCoV assay result |
|---|---|---|
| Ct < 37 | Any value | Positive. |
| 37 ≤ Ct < 40 | Any value | Inconclusive. Repeat the test.* |
| Ct = Undetermined or Ct = 40 | Ct < 40 | Negative. |
| Ct = Undetermined or Ct = 40 | Ct = Undetermined or Ct = 40 | Invalid.# |

* Once the test is repeated, the result for the assay is positive if Ct < 37 or if the result is consistent with the first test result of 37 ≤ Ct < 40.
### Re-purify the nucleic acid from the sample, then repeat the test.

Table 2. Interpretation of the SARS-CoV-2 test based on the Ct classification.

| 2019-nCoV assay result | Interpretation of results |
|---|---|
| Any two of the three assay are positive. | SARS-CoV-2 RNA is present. |
| Any one of the assays is positive in two different samples collected from the same subject. | SARS-CoV-2 RNA is present. |
| All three assays are negative. | SARS-CoV-2 RNA is NOT present. |

*Bacterial load analysis*
The bacterial load was assessed with Pan Bacterial 1 assay (Qiagen). The reaction was performed in a MicroAmp™ Optical 384-Well Reaction Plate (Thermo Fisher Scientific) in a QuantStudio™ 6 Flex Real-Time PCR System (Thermo Fisher Scientific) following manufacturer protocol. The bacterial load assessment was performed with DNA samples only. A standard curve was prepared using DNA extracted from 1 mL of a bacterial culture containing $9x10^7$ CFU/mL. The bacterial load was defined by comparing the resulted $C_t$ from Pan Bacterial 1 assay with the $C_t$ of from bacterial standard curve and adjusting by dilutions and volume used for extraction.
